# exRNA Signatures in Extracellular Vesicles and their Tumor-Lineage from Prostate Cancer

**DOI:** 10.1101/2020.09.28.20190009

**Authors:** Navneet Dogra, Mehmet E. Ahsen, Edgar EG Kozlova, Tzu-yi Chen, Kimaada Allette, Reena Olsen, Dan Han, Sung-Cheol Kim, Stacey M. Gifford, Joshua T. Smith, Benjamin H. Wunsch, Rachel Weil, Kamala Bhatt, Kamlesh K. Yadav, Konstantinos Vlachos, Sujit Nair, Ronald E. Gordon, Melissa Smith, Robert P. Sebra, Adam Margolin, Susmita Sahoo, Ashutosh K. Tewari, Carlos Cordon-Cardo, Bojan Losic, Gustavo Stolovitzky

## Abstract

Circulating extracellular vesicles (EVs) present in the bodily fluids of patients with cancer may provide non-invasive access to the tumor tissue. Yet, the transcriptomic lineage of tumor-derived EVs before and after tumor-resection remains poorly understood. Here, we established 60 total small RNA-sequencing profiles from 17 aggressive prostate cancer (PCa) patient’s tumor and adjacent normal tissue, and EVs isolated from urine, serum, and cancer cell culture media. We interrogated the key satellite alteration in tumor-derived EVs and found that resection of tumor prostate tissue leads to differential expression of reactive oxygen species (ROS), P53 pathways, inflammatory/cytokines, oncogenes, and tumor suppressor genes in the EV nanosatellites. Furthermore, we provide a set of novel EV-specific RNA signature, which are present in cancer but are nonexistent in post-resection patients with undetectable cancer. Finally, using a *de novo* RNAseq assembly followed by characterization of the small RNA landscape, we found novel small RNA clusters (smRCs) in the EVs, which reside in the unannotated regions. Novel smRCs were orthogonally validated for their differential expression in the ‘biomarker discovery’ cohort using RT-qPCR. We demonstrate that circulating tumor EVs provide a glimpse of the tumor tissue biology, resolving a major bottleneck in the current liquid biopsy efforts. Secretory vesicles appear to be playing a key role in non-canonical Wnt signaling and miRNA pathways, similar to the circulating tumor cells (CTCs), hence, we propose that such vesicles be called circulating tumor extracellular vesicles (CTEVs).

## Introduction

Extracellular vesicles (EVs) have brought tremendous excitement to the non-invasive liquid-biopsy procedures for detection, characterization, and monitoring of diseases^1-7^. EVs including microvesicles and exosomes are secreted by a variety of normal and tumor cells in bodily fluids^5, 8-14^. Hence, circulating EV “nanosatellites” contain molecular footprints (lipids, proteins, metabolites, RNA, and DNA) from their cell of origin^8*, 9, 15-20^. To date, numerous tissue-specific biomarkers have been discovered in the EVs derived from a variety of biofluids including saliva,^2, 15^ urine,^2, 12^ cerebrospinal fluid,^2, 21^ and blood^1-3^. Consequently, EVs are increasingly recognized as key players in the tumor microenvironment and metastasis, mainly through cellular crosstalk and vesicle trafficking, prompting their evaluation as prognostic and treatment response biomarkers^5, 11, 13, 22^. Although EVs show great promise for non-invasive diagnosis and monitoring of cancer, the transcriptomic lineage of tumor-derived EVs before and after tumor-resection remains poorly understood. This, in part, is due to the relatively small occurrence of tumor-derived EVs among all EVs accumulated from all cell types in biofluids. For instance, human blood contains ∼billion EVs per ml, which are accumulated from all tissue/cell types and only a fraction are originated from the tumor tissue of interest^23, 9, 12^. Furthermore, a significant clinical bottleneck is the unavailability of matched biopsied tumor and adjacent normal tissue, and bodily fluids from the same patients before and after cancer ^5, 13, 15, 24^. Hence, for the use of EVs cargo for diagnosis, one needs to show that they contain tissue-specific cargo, which alters (up- or down-regulated) during the disease progression and tumor resected cancer-free patients. In fact, little attention has been paid on the RNA of tumor-associated EVs once the tumor is removed, especially the comparison of small RNA cargo of circulating EVs before and after the patient is cancer-free.

Globally, Prostate cancer is the second-most common type of cancer^25, 26^. About 1.3 million US men undergo highly invasive prostate biopsy and ∼80% of such cases come out negative for prostate cancer annually^25, 27^. Prostate-Specific Antigen (PSA) is the current first line of evidence marker for PCa^26^. However, an increase in PSA levels can only indicate abnormalities in prostatic glands, but cannot clearly distinguish cancer.^26-29^ Hence, there is a need for detection accuracy and novel RNA-based liquid-biopsy biomarker discovery in prostate cancer. Prostate-specific EVs/exosomes (often called “prostatosomes”) are exclusively secreted by prostatic ductal epithelial cells into the seminal fluid, blood, urine, and other biofluids^12, 30^. Early EV-based liquid-biopsy studies are encouraging and provide new insight into the future of non-invasive, liquid-biopsy for cancer diagnostics^6, 30^.

In this context, we have investigated the key satellite alteration in the RNA signatures of tumor-derived EVs in cancer patients and monitored their occurrence as the tumor is resected and patients become cancer-free. We have conducted the total small RNA sequencing (n = 60) of aggressive prostate cancer patients (n = 17) tumor and adjacent normal tissue and EVs from urine, serum, and 22RV1 PCa cell line. We describe novel clinically-relevant tumor-associated RNA signatures in circulating EVs that can help detect and monitor cancer, which would allow patients to receive curative therapies. We also present a set of novel unannotated EVs specific small RNA clusters (smRCs), which resides in the largely uncharacterized noncoding regions of EVs RNA landscape and are present in prostate tissue, serum, and urine. We investigate key cancer mechanisms of reactive oxygen species (ROS), P53 pathways, inflammatory/cytokines, oncogenes, and tumor suppressor genes in the EV nanosatellites. Finally, our findings present a mini atlas of the intersection of tumor-derived exRNA in biofluids, enabling RNA signatures for liquid biopsy. To the best of our knowledge, this is the first study of its kind that has investigated the RNA lineage of EVs in the same patient at the time of cancer and 3-6 months after tumor resection once the patients are cancer-free.

## Results

Our study generated three independent prostate cancer RNA datasets (a total of 60 total small RNAseq from 17 aggressive prostate cancer patients tissue and EVs from biofluids): **1**) a “prostatic-lineage biomarker discovery” cohort, which includes tumor tissue, adjacent benign tissue, pre-prostatectomy serum, and urine derived Evs exRNA (total of 9 patients, 41 RNAseq); **2**) post-prostatectomy comparison “tumor exRNA discovery” cohort to identify differentially expressed exRNA between prostate cancer patients before and after they’re cancer free (n=8 patients x 2 (before and after prostatectomy) = 16 RNAseq); **3**) an independent “biomarker validation” cohort (n = 30 RT-qPCR) to confirm their clinical utility in a phase 2 biomarker study for detection of tumor-specific EVs RNA.

### Characterization of tumor tissue and biofluids-derived EVs

#### Characterization of vesicle secretion from tissue

To visualize the morphology and vesicle secretion from the prostate tissue, we investigated an aggressive PCa (Gleason score 9) patient’s tumor and adjacent benign tissue histology and electron microscopic imaging. First, the histology study confirmed the distribution of epithelial carcinoma throughout the tumor tissue sections (Fig. 1 c, e, h, and supplementary fig. 2b), which was entirely absent in the adjacent normal tissue (Fig. 1a, d, e and supplementary fig. 2a). Then, high resolution transmission electron microscopy (TEM) captured secretion of the vesicles into the lumen of prostate ductal epithelium from tumor and normal tissue (insets Fig. 1d, e). Electron micrographs of the tumor and normal tissue were morphologically unique. While the prostatic glands were intact in normal tissue, carcinoma of the prostate is observed in the luminal epithelial cells of the tumor. An important observation was that a relatively higher number (∼2.6 X) (Fig. 1g) of vacuoles, vesicles, and multivesicular bodies (MVBs) were present in the tumor epithelium (∼400 MVBs were measured from 3 different tissue sections, supplementary methods). In fact, elevated levels of EVs have previously been reported in ovarian and PCa patients in biofluids^6, 23^. These studies were conducted with EVs isolated from respective biofluids using a nanoparticle tracking analyses (NTA) system, which measures Brownian motion of the colloidal particles^23^. However, electron microscopic visualization and quantification of secretory vesicles has not been reported yet^6, 23^. We found that over ∼90% of the enlarged vacuoles and MVBs were present in the luminal prostate ductal epithelial carcinoma, while no major changes were observed in the stroma of the tumor tissue. We observed two unique mechanisms of EVs secretion, (**1**) vesicles of endocytic origin secreted via fusion of MVBs with the plasma membrane (Fig. 1e inset), (**2**) secretion via budding (exocytosis) from the plasma membrane (Fig. 1d inset). Notably, the first mechanism displayed consistent round and cup-shaped ∼80nm vesicles secretion, while second mechanism displayed relatively larger and heterogenous, ∼50-250 nm vesicles.

**Fig. 1.**
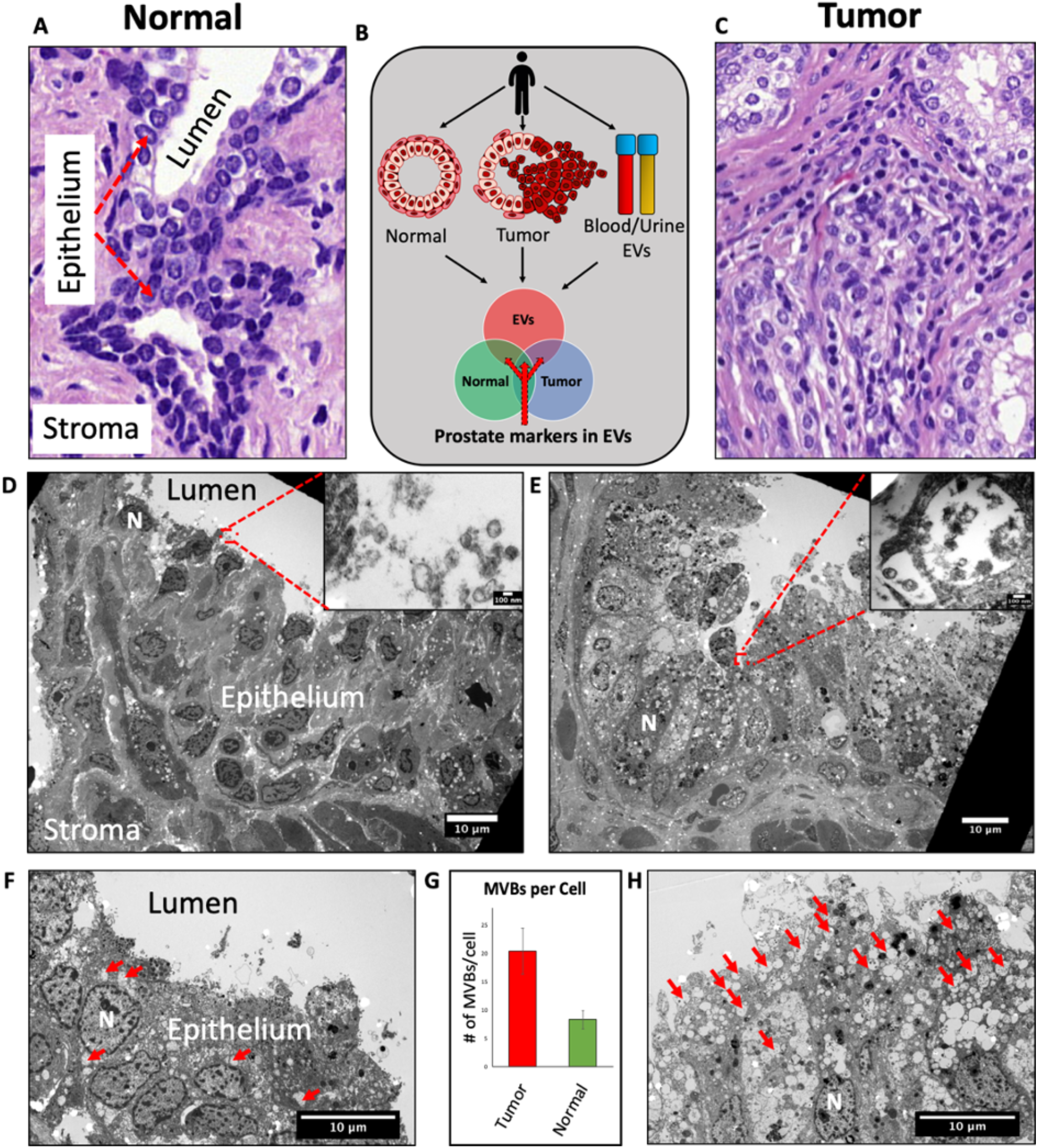
A comparison between prostate cancer patient’s normal and tumor tissue revealed different morphology, higher number of MVBs, and vesicles secretion. Histological analyses of a ∼70-75 year old PCa patient (Gleason score 9) normal (A) and tumor (C) tissue. Transmission electron microscopic analyses of the same PCa patients’ normal (D) and tumor (E) tissue (Inset. vesicle secretion in the luminal area of prostate tissue). (B) A schematic of the overall design of present study, where patient-matched prostate tumor, normal, and bodily fluids are studied. High resolution images of epithelium in normal (F) and tumor (H) tissue, where higher number of MVBs can be clearly seen in tumor (MVBs are marked red, N is marked as nucleus of the epithelial cells). (G) Quantification of multivesicular bodies (MVBs) showed that relatively higher number (2.6 times) were present in the ductal epithelial carcinoma of the prostate. Scale bar for Fig. 1B & 1C are 10um (Inset scale bar is 100 nm), 1E = 100 nm.

#### Characterization of isolated vesicles

We next isolated EVs from patient serum, urine, and cell culture medium using either UC and/or nanoDLD device. Our nanoDLD vs UC comparison of the EVs exRNA was recently published in a major NIH Extracellular RNA Communication Consortium (ERCC) study^2^, which investigated and compared exRNA cargo types in over 5000 human samples using different isolation methods and biofluids. This independent analysis found that our isolation methods specifically isolate low- and high-density vesicles (cargo type 1 & 4, respectively) with minimum contamination from the lipoproteins and argonaute proteins^2^. Here, we conducted additional quality control of the EVs, which included TEM, immuno-gold TEM, NTA, immuno-fluorescence labeling, and Exoview™ (for canonical transmembrane tetraspanins). The isolated EVs from serum, urine, and cell culture media were observed in the range of ∼50-200 nm diameter in a typical cup-shaped morphology, similar to the ones seen secreted from tissue (Figure 2a and supplementary fig. 3A-C). To confirm the presence of canonical tetraspanins in the EVs, we conducted an immuno-gold TEM (with 6 nm gold - CD81 antibody), showing enrichment of CD81 protein on the surface of EVs (Fig. 2b). Additionally, multi-color immuno-fluorescence co-localization of EVs functionalized on a substrate confirmed the presence of three major canonical tetraspanin EV markers CD81, CD9, and CD63 on their surface (Fig. 2c). Mouse IgG was non-specific to EVs and displayed no significant signal for exosomal tetraspanin proteins. Of note, the concentration of serum EVs was ∼1000x higher (∼10^12^ particles/ml) (Fig. 1F) than of EVs from urine and cell culture media (10^9^ particles/ml) (supplementary fig. 3A-C). Variable amounts of EVs may be dependent upon the fluids type and the volume used for EVs isolation. For instance, EVs were extracted from 2ml of serum, 25ml of urine, and 200ml of cell culture media. Regardless of the source of biofluid, all EVs yielded zeta potential ranging between −5 to −35 mV, indicating their incipient to moderate stability (supplementary fig. 3A-C).

**Fig. 2.**
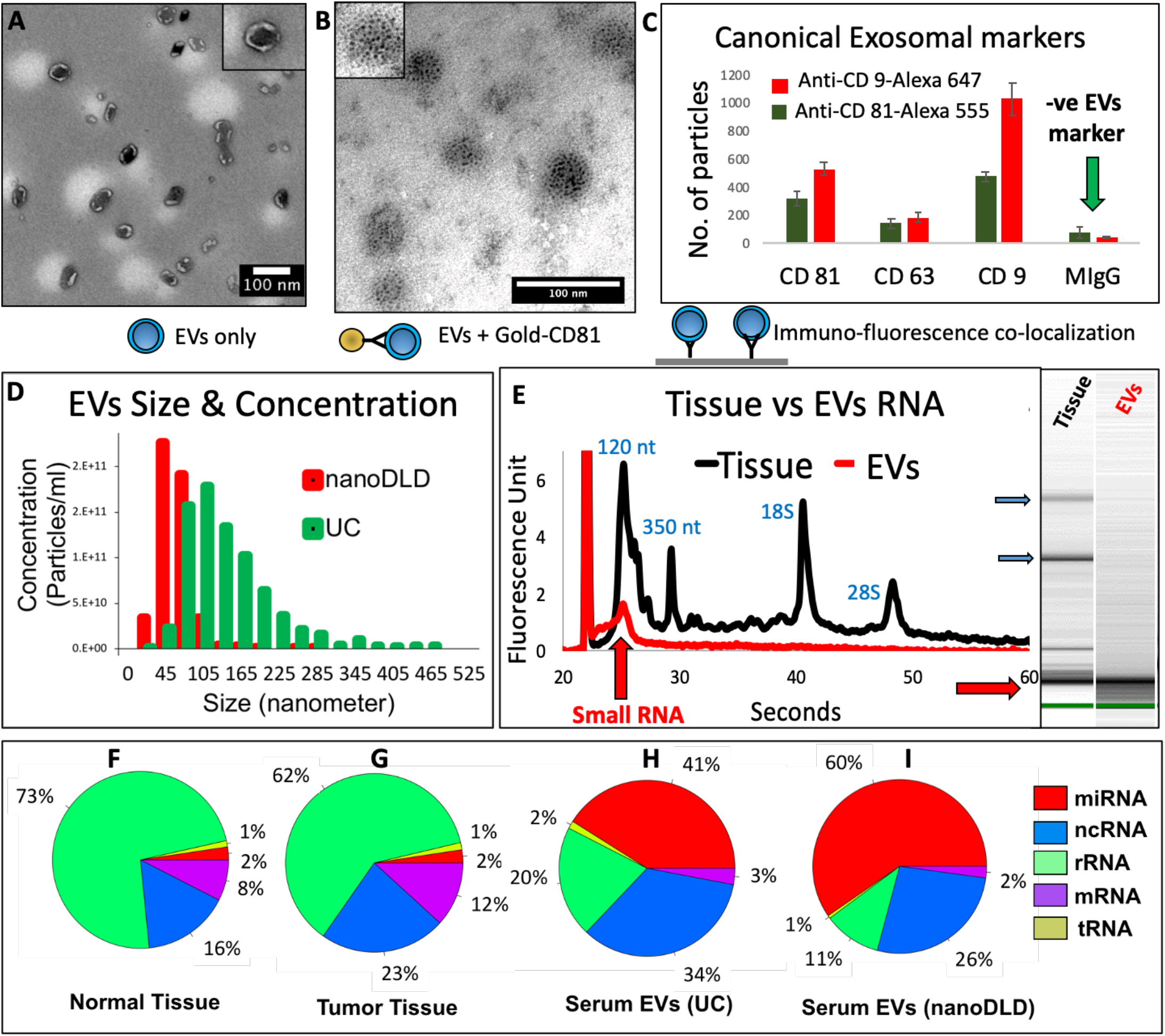
Serum and urine derived EVs primarily contain small non-coding RNA (∼40-60% of total RNA was miRNA), while tissue predominantly yielded rRNA (>60%). (**A**) TEM image of serum EVs isolated using nanoDLD chip (**B**) canonical markers Immunogold (6nm) labeled CD81 shows specificity to the vesicle surface. (**C**) nanoview analyses of nanoDLD isolated EVs confirmed presence of canonical tetraspanins on vesicle surface. Mouse IgG antibody is a negative control. (**D**) A size and concentration comparison of nanoDLD versus UC isolated EVs. nanoDLD EVs were of relatively narrower distribution (50-100 nm), while UC exhibit broader distribution of vesicles (50-250 nm). (**E**) Electropherogram comparison between tissue RNA (black curve) and serum EVs RNA (red curve) exhibit that EVs primarily contained small RNA (lower red arrow in the gel) and did not display ribosomal RNA (top blue arrows). Small RNA is marked 120 nt (nucleotide) and 350 nt. Large ribosomal RNA are marked 18S and 28S. Post sequencing quantification of RNA types yielded normal (**F**) and tumor tissue (**G**) primarily contained rRNA, while serum (**H** and **I**) EVs primarily contained miRNA and other ncRNA.

#### Characterization of vesicular RNA

Next, we asked if the size distribution of RNA that is packed inside the vesicles are unique among tissue and EVs. To address this, we conducted capillary electrophoresis from cancer patient’s tumor and adjacent benign tissue, and EVs from serum, urine, and cancer cell culture. As presented in recent publications^8, 9^, EVs electropherograms exclusively displayed small RNA (<200 nt), while the tissue displayed both the small RNA and large ribosomal RNA (rRNA, 18S & 28S) (Fig. 2e, Supplementary Fig. 4). Although EVs isolated from serum, urine or cell culture media displayed subtle differences in the electropherograms, all EVs primarily contained small RNA cargo (Supplementary Fig. 4). Then, to interrogate the distinct RNA subtypes in tissue vs EVs, we conducted a total small RNA sequencing of all specimen (n = 60). The total small RNA sequencing revealed that non-coding RNA (ncRNA) are the major source from EVs, yielding as much as ∼70-87% of ncRNA in the vesicles (Fig 2h-i). When studied among RNA subtypes, miRNA was the major source from EVs (∼40-60%), while only a small percentage of mRNA (∼2-3%) and tRNA (∼1%) were present in EVs (Fig. 2h-i). In contrast, tissue displayed ∼60-70% of rRNA, while only showing ∼20% of ncRNA (Fig. 2f, g). The amount of RNA subtypes in vesicles is important, as recent findings have reported conflicting amounts (40-90%) of ribosomal RNA (rRNA) in exosomes and other EVs. Consequently, there remains an ongoing debate regarding the presence (or absence) of ribosomal RNA in the EVs^18, 31^. In our study, we found extremely small amounts of rRNA in the EVs (∼11-20%) (Fig. 2f, g). We found that the measured RNA was dependent on the technology used for EVs isolation (UC and nanoDLD). Of note, nanoDLD device was designed to enrich for ∼70 nm particles, precisely excluding smaller (<50 nm) and larger particles (>150 nm). As a result, nanoDLD yielded relatively homogeneous ∼50-80nm vesicles, while UC isolated larger heterogeneous particles (∼50-250 nm) (Fig. 2d). The resulting size-based differences in vesicles were also translated into the post RNA sequencing outcome of UC vs nanoDLD isolation. In comparison with UC, nanoDLD isolated vesicles revealed higher miRNA (∼60% vs 41%) and lower rRNA (11% vs 20%), respectively. These results indicate that the smaller (∼70 nm) vesicles primarily contain small ncRNA and most of the rRNA contamination may have been contributed from the larger apoptotic and budded vesicles, which may not be of endocytic origin. Based on these findings, we hypothesize that either larger vesicles (>150 nm) were the carriers for rRNA, or membrane-free rRNA was pelleted at the same sedimentation velocity as EVs during the UC isolation.

### Prostate tissue-specific gene lineage is present in the circulating EVs, which downregulated post-resection

We asked whether the prostate-specific RNA signatures exist in biofluids-derived EVs and if such biomarkers can be reproducibly studied for liquid biopsy. To address this, we employed 3 different strategies. **1**) A literature curated prostatic gene-set; **2**) Intersection of RNAseq amongst prostate tissue and EVs to identify overlap; **3**) Differential expression of RNA signatures from pre- and post-tumor resection EVs.

#### 1) Literature curated prostatic markers and their presence in the Evs

To test whether the prostate-specific RNA signatures are present in the circulating EVs, we curated a set of 22 vastly studied and published RNA markers, which are either prostate-specific or cancer-specific (Supplementary Table. 2)^26^. For instance, PSA-encoding kallikreins (KLK2, KLK3, and KLK4), androgen receptor (AR), and NKX3-1 are exclusively expressed in the prostate tissue (curated from 53 tissue/cell type in GTEX portal, Supplementary Fig. 7a-d)^32^. In addition, ncRNAs such as MALAT1, NEAT1, MIR25, and LET-7 are associated with PCa^6, 12, 27, 33, 34, 35, 36, 37^. Moreover, prostate tissue is comprised of epithelial (EPCAM, EGFR, KRT8), basal (BCAM), and stromal cell (CD44, CD105, CD29) RNA lineage^38, 39^. Hence, combining all prostate tissue-specific markers yielded an overall prostate tissue lineage gene set (Fig. 3a, supplementary Fig. 6a). In contrast, endothelial (ICAM1, MCAM, Sele) and leucocyte markers from hematopoietic RNA lineage (CD45, CD16, CD41), are known to be absent in prostatic tissue and are used as negative prostatic markers (Fig. 3A) in our study. Similar RNA lineage studies have previously been conducted for CTCs in blood^38, 39^.

**Fig. 3.**
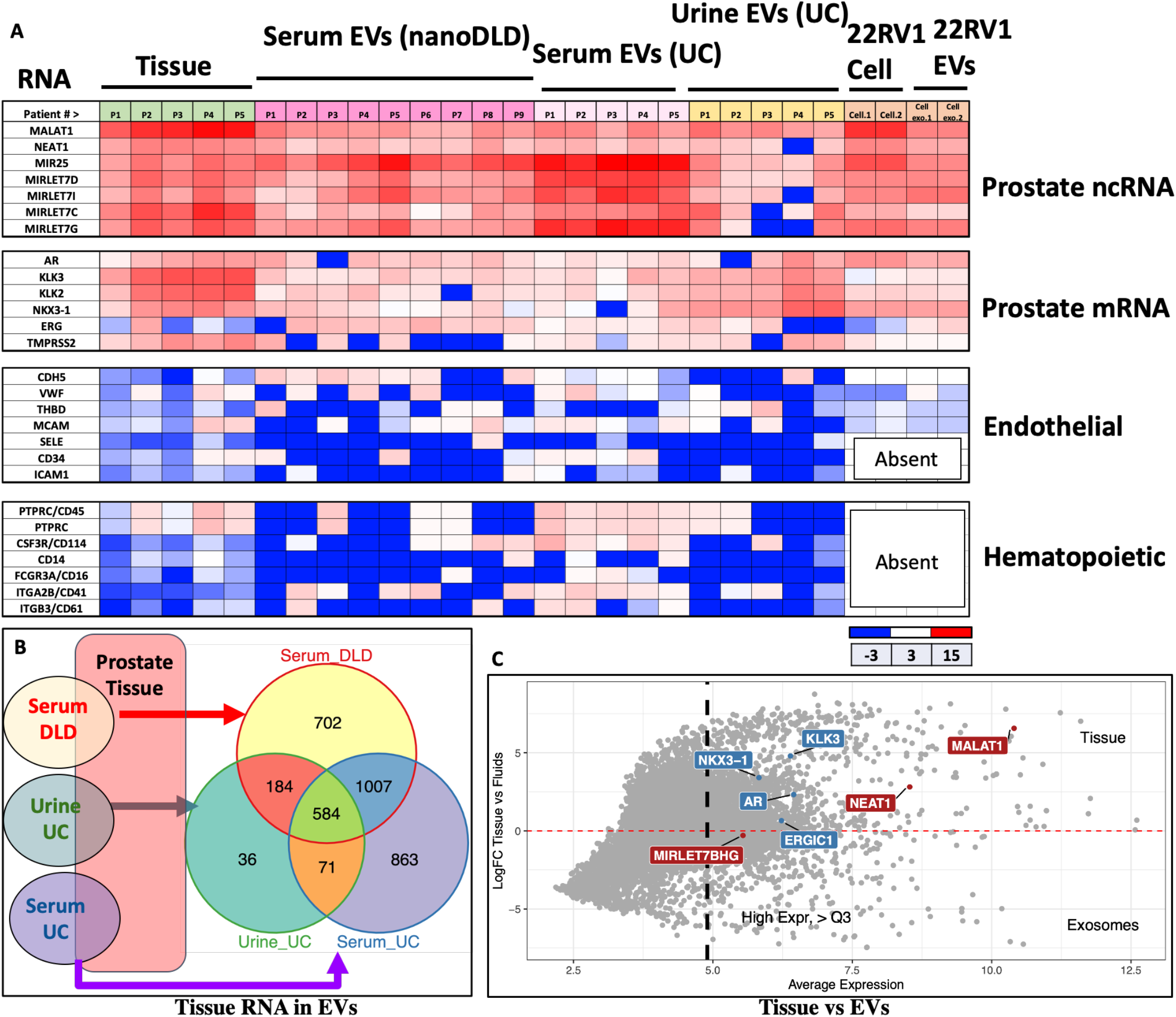
Prostate tissue-specific gene signatures are present in serum, urine, and cell culture derived EVs. (**A**) High resolution heat map showing expression of literature curated gene sets in PCa tissue, nanoDLD isolated serum EVs, UC isolated serum EVs, UC isolated urine EVs, PCa cell line, and cell line derived EVs. Prostate coding and noncoding markers are highly expressed in EVs, confirming that EVs contain prostate markers. Non-prostatic endothelial and hematopoietic markers were negative controls and do not show high expression for prostate tumor tissue and EVs. (**B**) Overlap between prostate tissue RNA and serum/urine EVs RNA. Final analyses present a set of common RNA present in prostate tissue and serum/urine EVs. (**C**) Average expression of tissue RNA versus EVs RNA in biofluids.

First, the unsupervised hierarchical clustering analysis of all KLK genes resulted in distinct clusters of prostatic KLKs (KLK3, KLK2, and KLK4) in tissue, showing an expected abundance of the prostate tissue. (Supplementary Fig. 6b). In contrast, hematopoietic and endothelial RNA markers were absent in the tissue, EVs, and the PCa cell line (Fig. 3a), confirming the absence of leucocyte and platelet (CD16, CD41) contaminants in the EVs. Next, we compared a set of prostatic tissue lineage genes set with the RNA discovered in EVs from different fluids (serum, urine, and cell culture medium) (Fig. 3a). Indeed, prostate-specific RNA markers were expressed in prostate tissue and EVs, consistent with our hypothesis of the presence of prostate derived EVs in biofluids outside the prostate (Fig. 3a). Interestingly, certain prostatic RNA yielded higher expression in urine EVs, while others were upregulated in the serum EVs. For instance, higher expression of NKX3-1, KLK2, KLK3, and AR in urine EVs is possibly due to the proximity of prostate to the urinary tract^27^.

#### 2) RNA Intersection amongst prostate tissue and EVs and their molecular pathways

In order to identify circulating tissue-specific EVs biomarker, such satellite RNA molecule must overlap both in the tissue and in EVs of cancer patients. We conducted a detailed analysis of the intersection RNA, which yielded a set of 584 common RNA markers overlapped in prostate tissue, serum, and urine derived EVs from 9 aggressive cancer patients (Fig. 3b). Out of the ∼15,728 RNA transcripts studied in prostate tissue, only 875 (5.5 %) were common amongst urine EVs (Fig. 3b). Similar analyses on serum UC and nanoDLD EVs yielded ∼16 % of prostatic RNA in the EVs (Fig. 3b and Supplementary Fig. 8). In addition, we discovered a set of upregulated tumor markers present in PCa patient’s EVs (Fig. 3c). Some EV RNA such as Testis-Specific Kinase 1 (TESK2), Spondin 2 (SPON2), MNX1, CANT1, PRSS36, ABCC4, and TMEFF2 were among the highly upregulated genes in EVs. SPON2 is previously proposed as a diagnostic candidate marker for prostate.^40^ MNX1 has been suggested as a new oncogene in prostate cancer,^41^ while ABCC4, CANT1, and TMEFF2 are androgen-regulated genes closely related to PCa progression.^42, 43, 44^ Although, TESK2 was upregulated, it has not yet been studied for prostate cancer. Among the 53 tested tissue types, TESK2 is the highest expressed genes in prostate tissue (Supplementary Fig. 7f) and high expressions of TESK2 in prostate and its release in EVs may enable a novel marker to noninvasively monitor prostatic tumorigenesis. Next, to determine the association between patients’ tissue and EV profiles, we measured pairwise correlation of the RNAseq of tumor and adjacent normal tissue, and EVs from serum, urine. All specimen RNAseq clustered in their respective categories (Fig. 4b). Serum EVs clustered in two subgroups, where nanoDLD EVs remained together and UC EVs clustered together in a subgroup (all serum specimen clustered in same group) (Fig. 4b). Then, we attempted to gain information about the similarities of RNA between tumor and adjacent normal tissue from PCa patients, which yielded a Rho = 0.97, indicating that only small transcriptomic changes induce striking differences among the tissue (Fig. 4a). Correlation between serum and urine EVs RNA from the same patients yielded Rho = 0.36, which is consistent with our hypothesis that urine and serum EVs cargo unique RNA, essentially owing to the proximity of their tissue of origin (Fig. 4a). In contrast, correlation between tumor versus serum EVs yielded Rho = −0.18 (Fig. 4a), suggesting that most abundant transcripts in tissue specimen were different than in the EVs, reinforcing our hypothesis of vesicles accumulation from all cell types and disposal/delivery mechanism from EVs. Previously, similar studies in cell lines and their EVs have yielded low correlation (R^2^ = 0.01), confirming that regardless of the tissue or cell line of origin, EVs carry a unique set of RNA.^8^

**Figure 4.**
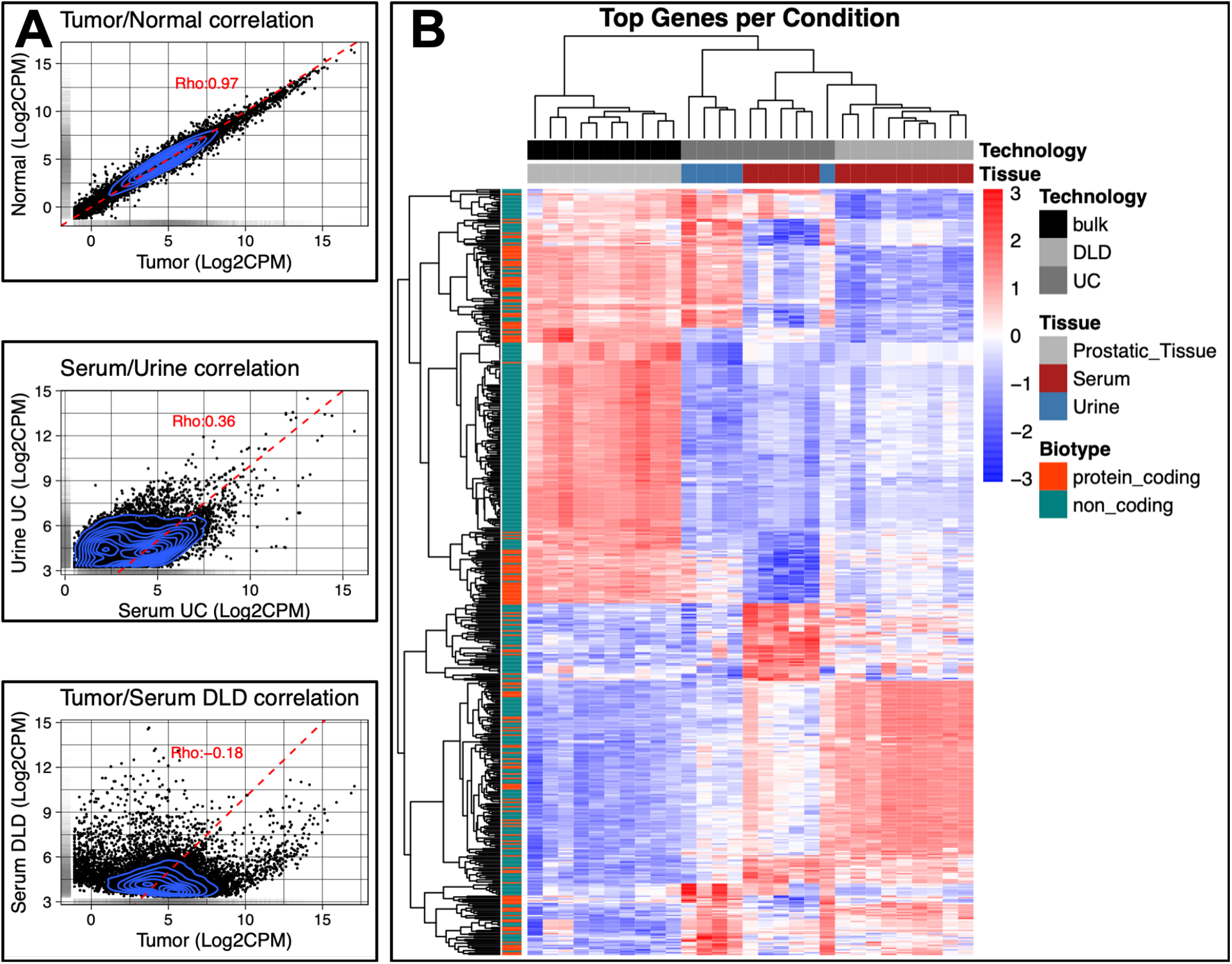
A Correlations between tissue and EVs. (**A**) Correlation between Tumor/Normal, EVs isolated from SerumUC/UrineUC and Tumor versus Serum EVs. (**B**) Heatmap with top 500 differentially expressed Loci between comparisons, hierarchically sorted.

#### EVs mainly enrich for signaling pathways

In order to investigate the key molecular pathways packaged in EVs, we conducted pathway enrichment analyses using three different databases KEGG, wikipathways, and NCI-Nature^45, 46, 47, 48, 49^. In total, over 30 major pathways (P ≥ 0.01) were enriched in all EVs. A major finding was that EVs overwhelmingly (>90%) enriched for miRNA, T-cell receptor, hormonal, cytokines, and growth factor related pathways, all of which are signaling pathways (Fig. 5a)^38^. EVs ncRNA LET-7D/ -G/ -I, and MIR25 are known to play major role in tumorigenesis and angiogenesis, and their presence in EVs, may play a key role in seeding and dissemination of cancer^8, 36, 50, 51^. Primarily miRNA and Wnt signaling pathways were enriched in the EVs and non-canonical Wnt Signaling (P = 0.0039) was enriched in ∼80% (14/17 specimen) of the PCa patient serum EVs, implicating circulating EVs role in Wnt signaling associated seeding and dissemination mechanism (Fig. 5c, Supplementary Fig. 9). Recent studies suggested that EVs are capable of inducing Wnt signaling in recipient cells, indicating that EVs derived Wnt pathways may regulate key cellular processes in cell proliferation, polarity, migration, stem cell renewal, and apoptosis^52, 53, 52^. SERPINE1 and FN1 gene signatures were abundant in EVs and are implicated in epithelial to mesenchymal transition (EMT) (Supplementary Fig. 6a), indicating that EVs may play a key role in EMT in cancer, which remain to be studied in details^54, 52, 53^. Of note, androgen receptor signaling pathway (Adj. P value = 0.09) was among the top 5 pathways in prostatic EVs RNA. AR pathway is extensively studied in prostate cancer, and its traces found in circulating EVs may enable non-invasive monitoring of hormone associated diseases.

**Fig. 5.**
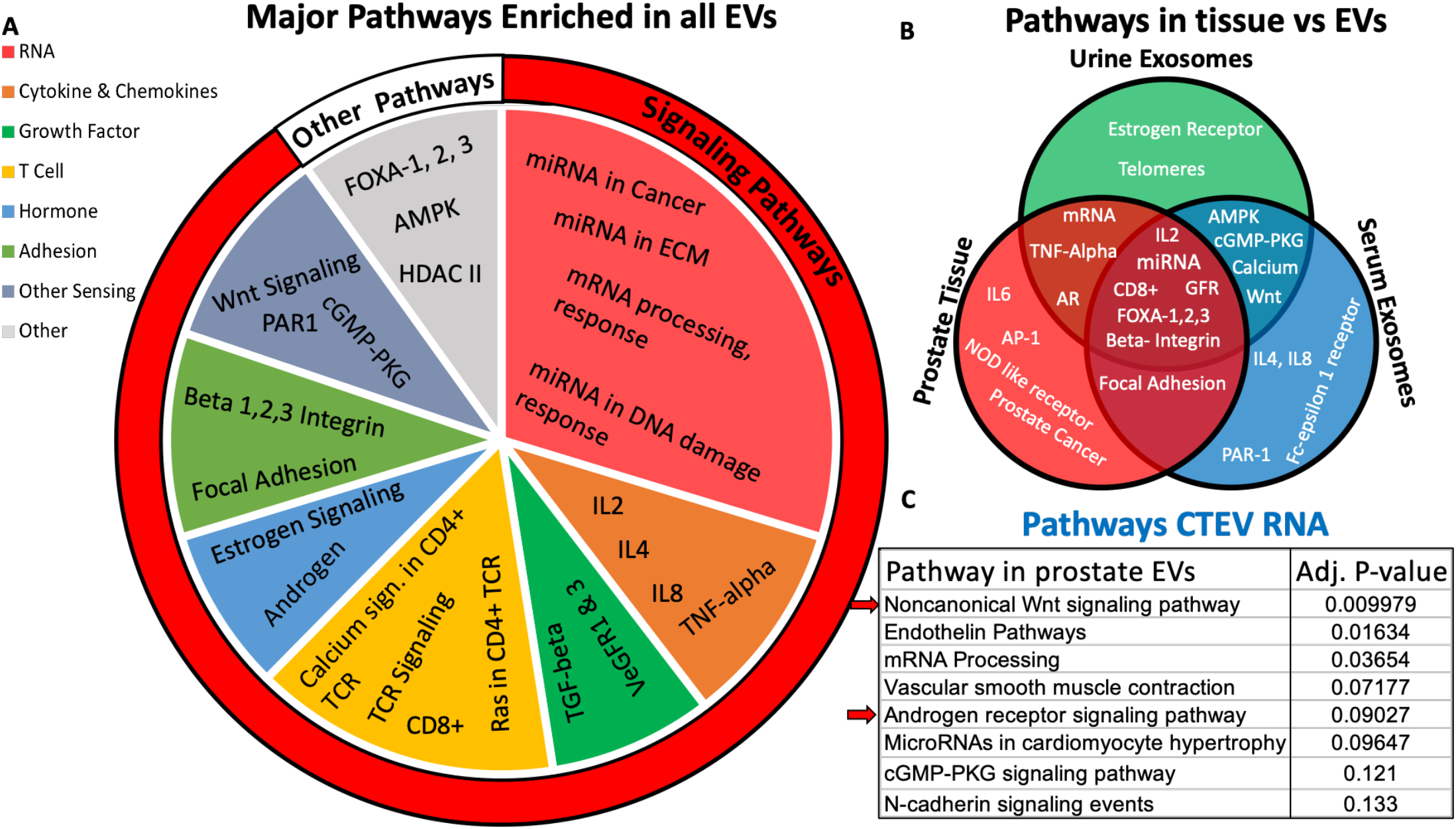
Serum and urine EVs primarily (>90%) enrich for signaling associated pathways, while prostate-specific EVs RNA contribute to major hormonal pathways. Three separate databases (KEGG, Wikipathways, NCI-Nature) were used for pathway enrichment study. (**A**) EVs were primarily (>90%) enriched for signaling pathways, showing miRNA, cytokines, hormone, T-cell receptor (CD8+, CD4+), growth factor, and adhesion pathways. (**B**) Tissue, serum EVs, and urine EVs were enriched for certain overlapping and some unique pathways. (**C**) Prostatic CTEV RNA yielded unique pathways and primarily enriched for prostate-related hormonal androgen receptor pathways, Wnt signaling, endothelin, and mRNA processing are some highly studied pathways in prostate.

#### 3. Differential expression of RNA signature pre- and post-tumor resection

In order to examine the overall genomic landscape of cancer RNA in the EVs, we leveraged three different data sets for generating a cumulative group of most significant gene-sets in cancer and their manifestation in the EVs. **1**) COSMIC cancer database - curated data from large scale experimental screens from the Cancer Genome Project at the Sanger Institute. **2**) Molecular Signatures Database (MSigDB) - annotated gene sets for use with GSEA software, and **3**) Literature curated Tumor suppressor genes, oncogenes, and cytokines. Below we discuss in-depth detail of major cancer pathways in before and after tumor-resection cancer patients.

### Cytokines, Oncogenes, and tumor suppressor genes are differentially expressed pre- & post-tumor resection EVs

Cytokines, oncogenes and tumor suppressor genes are widely studied in cancer, however, their presence in EVs from pre- and post-resected cancer patients has not been studied yet. We asked whether the cancer derived EV-RNA (pre-resection) differ from the cancer free patients (3-6 months post-resection, undetectable PSA). To test for the differential expression of EV-RNA pre- and post-resection, we computed the fold change and false discovery rate. We found the many of the upregulated genes in cancer EVs were also some the most frequently studied oncogenes and were upregulated in cancer than those in the cancer free patients (Fig. 6a). One of the most interesting finding was that major tumor suppressor genes (NKX3.1, BRCA1, MDX4) were highly expressed in the cancer patient EVs (Fig. 6a, c)^55^. This is counter intuitive, as tumor suppressor genes tend to be downregulated in tumor tissue^33, 55^. For instance, NKX3.1 is androgen regulated tumor suppressor genes, which is predominantly localized in the prostate epithelium^33^. BRCA1 interacts with estrogen receptors and its expression is downregulated and becomes tumorigenic in high grade breast, ovarian, and other cancers^25^. As it is widely known that the deletion of expression of NKX3.1 and BRCA1 correlates with tumor progression, one may also expect their lower expression in the circulating EVs^33^. In addition, resection of tumor resulted in downregulated ROS, P53 pathways, inflammatory cytokines, and major oncogenes in EV nanosatellites (Fig. 6b). The overexpression of the tumor suppressor genes and inflammatory cytokines in the EVs of cancer patients can be explained by following mechanisms: **1**) The disposal/delivery mechanism from EVs may be playing a key role in the dissemination of tumor-associated (oncogenes or tumor suppressors) RNA. **2**) The overexpression of cytokines in the EVs may stem from the immunological responses of the EVs. Remarkably, these results also support the hypothesis that upregulated cancer circulating EVs, which downregulates in cancer free patients may play a key role in the dissemination of tumor-associated RNA and may be viable target for liquid biopsy biomarkers.

**Figure 6.**
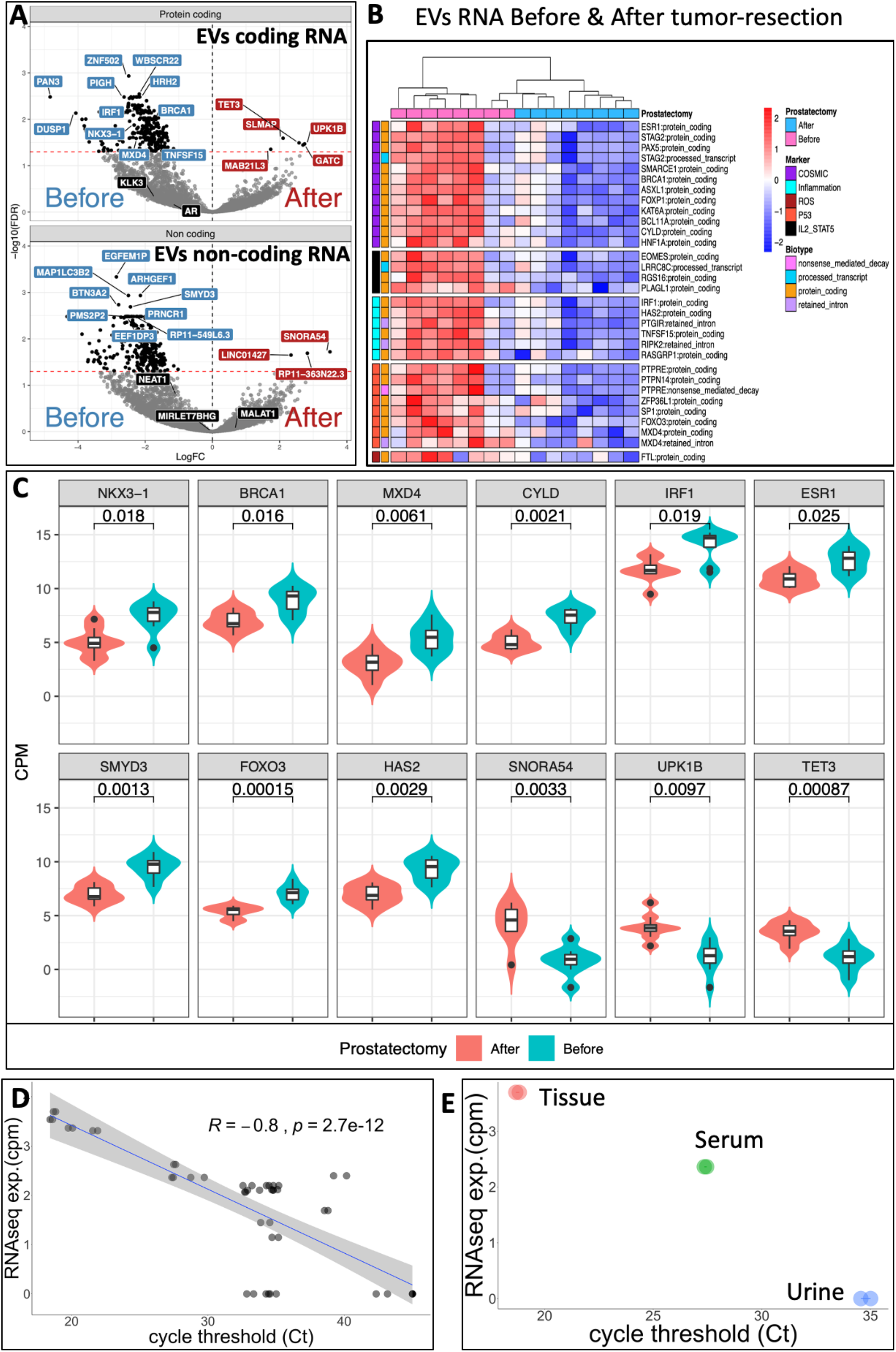
Differentially expressed RNA in pre- & post-tumor resection in cancer free patients. (**A**) Volcano plot of before and after tumor-resection showing coding (top) noncoding (bottom) RNAs. (**B**) Heatmap showing significant locus labeled by known curated markers from COSMIC and Hallmark database for cancer, inflammation, ROS, P53 and IL2/STAT5 pathways. (**C**) Major differentially expressed RNA, which included tumor suppressor genes, oncogenes and miRNAs. (**D**) The higher RNAseq expression yields lower Ct for novel unannotated RNA biomarkers (smRCs), which were orthogonally validated using step-loop RT, followed by qPCR. (**E**) Differential expression of smRCs in tissue, serum, and urine correlated with RNAseq expression.

**Figure 7.**
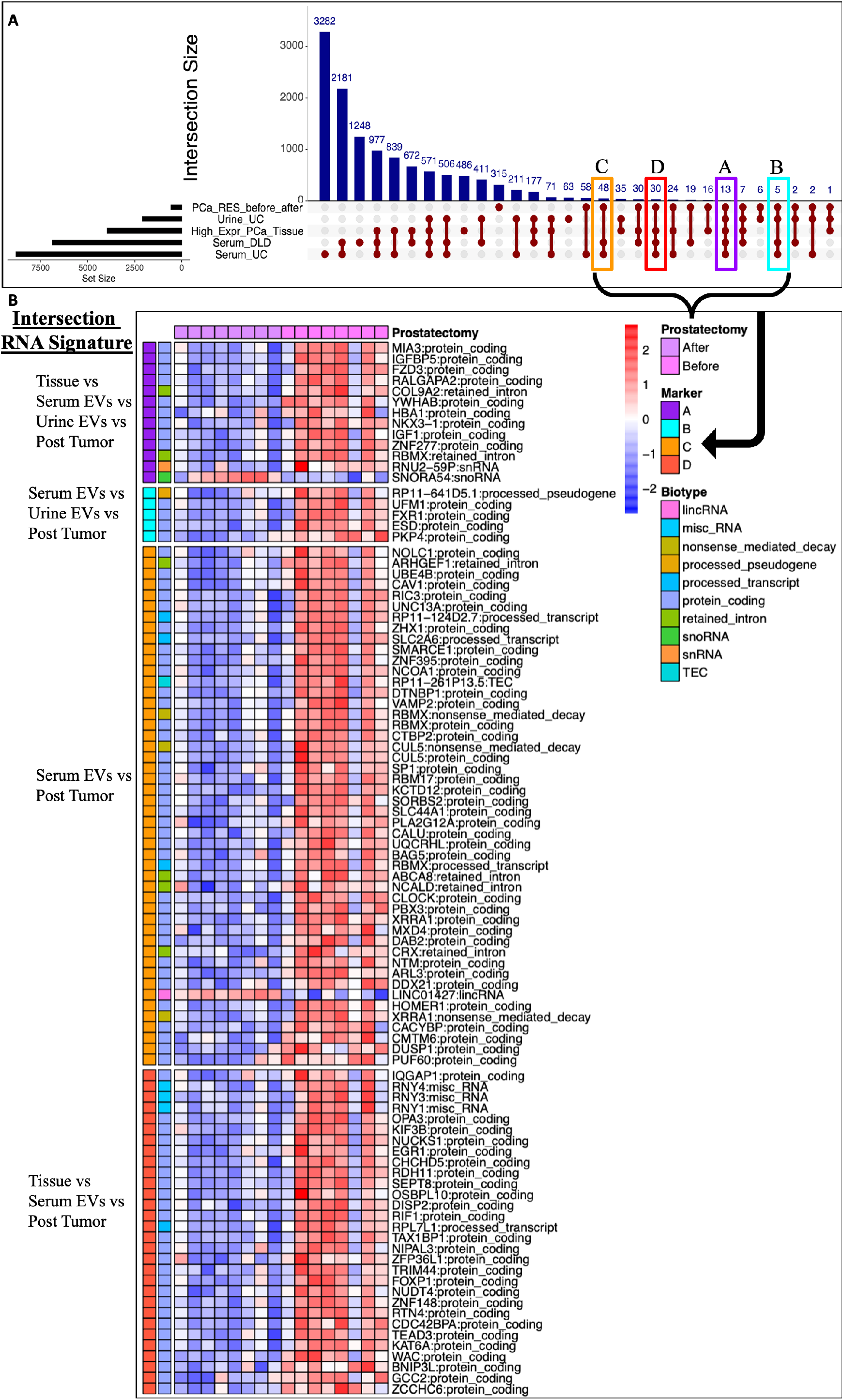
Mini Atlas of RNA signature that intersect among different biofluids and tissue. **(A)** An upset plot showing the locus common to each intersection of signatures (pre-/post-resection, urine, serum and high expression in tissue). (**B**) Major intersections in EVs RNA signatures across tissue and fluids, which will hopefully enable noninvasive monitoring of cancer biomarkers, identify tumor specific RNA, and may benefit by avoiding unnecessary surgical procedures.

### EVs cargo novel small RNA clusters (smRCs)

Most RNA-based studies apply conventional reference based approach and thus are limited to the annotated genomic regions. This approach is sufficient for cell-based analyses, however, since EVs predominantly contain small noncoding RNA, it becomes apparent that small EV-RNA requires a different view for a better understanding of these nanosatellites. To identify the unannotated small RNA that are present in the EVs, we conducted a *de novo* assembly followed by characterization of the small RNA landscape of the unannotated regions. Given their high variability among tissue and fluid types, we computed tissue specificity of smRCs and traced them in the biofluids. We selected the three top smRCs (see Supplementary Methods) and orthogonally validated their differential expression in our ‘biomarker discovery’ cohort using stem-loop RT-qPCR. In fact, the RNAseq results were highly correlated with RT-qPCR outcome, discarding our hypothesis of alignment or other computational bias (Fig. 6d). The Pearson’s correlation coefficient was higher than 0.8 for all three smRCs when comparing data from small RNA sequencing and RT-qPCR (p = 2.7e^-12^, Fig. 6d). The three smRCs were located in regions of chromosomes 2q, 8q (chr8:21329709-21329879, chr2:148881489-148881928, chr2:222918489-222918711). Interestingly, some of the smRCs in our *de novo* RNAseq analyses were selectively present in serum EVs but were absent in urine EVs. This allowed us to validate them with RT-PCR and identify biofluid-specific smRCs for selective fluid-based diagnostics (Fig. 6e). Taken together, our analyses show that EVs unannotated RNAs that arise from endogenous genes and are part of the genomic ‘dark matter’ of highly abundant yet largely uncharacterized non-coding RNA, may play a key emerging role in regulating gene expression, post-transcriptional, and translational mechanisms.

## Discussion

We interrogated the key satellite alteration in tumor-derived EVs and found that resection of tumor prostate cancer tissue leads to differential expression of novel RNA biomarkers, ROS, P53 pathways, inflammatory/cytokines, major oncogenes, and tumor suppressors in the EV nanosatellites. Perhaps our most unexpected finding is that the tumor suppressor genes were upregulate in EVs. A key remaining question is how does NKX3.1, BRCA1 and oncogenes’ RNA is disseminated from tumor to the biofluids. Regardless, these findings contain major diagnostic implications for cancers with precise measurements of up- or down-regulation of key tumor-specific EV-associated RNA that we put forth in this manuscript. Furthermore, we present a mini atlas based on our study of independently generated and cross-validated three prostate cancer EV-RNA datasets (a total of 60 RNAseq from 17 aggressive prostate cancer patients). This dataset provides detailed RNA signatures, which may lead to substantial future discoveries in EV-based liquid-biopsy. Below, we discuss in-depth and speculate as to how tumor-derived EVs may contribute to cancer progression and provide next-generation fluid-based monitoring of tumor-derived RNA, a key barrier in non-invasive cancer diagnostics.

### Could EVs play a role in the pathogenesis of cancer?

EVs may have a hitherto unknown role in cancer as a regulator of microenvironment, a metabolic adaptation employed by cancer cells. Importantly, our exRNA-derived EV signature can be developed as a method for cancer surveillance and not just as a diagnostic tool.

### Could EVs RNA contribute to liquid-biopsy in cancers?

The implications of our findings extend beyond prostate cancer. As ROS, cytokines, P53 has implications in many cancers and non-cancers, we speculate that the EVs RNA are relevant to other diseases.

### Could EVs play a role in the therapeutics of cancer?

It is important to mention that in our studies urine and serum were enriched in many distinct pathways, revealing that based on the source of biofluid of EVs, unique RNA can be isolated, enriched, and studied. This outcome has direct implication in therapy.

In summary, we have investigated patient matched tumor, adjacent normal tissue, and EVs from serum, urine, and cell culture in prostate cancer. We demonstrate that EVs are predominantly enriched in small ncRNA, encapsulating ∼40-60% of miRNA. Gene set enrichment analyses revealed that EVs are primarily (>90%) enriched in signaling pathways. Finally, we show that resection of tumor prostate tissue leads to differential expression of ROS, P53 pathways, inflammatory/cytokines, major oncogenes, and tumor suppressors in the EV nanosatellites. Future studies along the lines described here will hopefully enable noninvasive monitoring of cancer biomarkers, identify tumor specific RNA, and may benefit by avoiding unnecessary surgical procedures.

## Data Availability

The RNAseq data is published on exRNA atlas https://exrna-atlas.org/.
The data will also be available on GEO after publication.

https://exrna.org/

